# Empirical contact networks reveal heterogeneous outbreak risks in a UK long-term care facility: a modelling study

**DOI:** 10.64898/2026.07.07.26357238

**Authors:** Li Pi, Emma L. Davis, Leon Danon, T. Déirdre Hollingsworth

## Abstract

Long-term care facilities (LTCs) worldwide experienced disproportionately high infection and mortality rates during the COVID-19 pandemic, where essential care limits opportunities for contact segregation. However, empirical contact data remain scarce, limiting our understanding of how individual contact behaviours shape transmission in these settings. In this study, we developed a stochastic network-based transmission model parameterised using real-world self-reported contact data collected from a median-sized UK LTC unit. By incorporating high-resolution observational data that reflect routine care delivery patterns, we quantified how heterogeneity in contact networks influences outbreak dynamics.

We found substantial variation in contact behaviour between individuals, resulting in highly heterogeneous transmission outcomes. Outbreak occurrence, timing, final size, and the likelihood of super-spreading events all varied markedly depending on the structure of the underlying contact network and the characteristics of the index case. Individuals with high contact activity were considerably more likely to initiate large outbreaks than those with fewer contacts. For a per-contact transmission probability of 10%, introduction of infection through the most highly connected individuals resulted in a greater than 75% probability of a large outbreak.

Our findings indicate that preventing infection introduction through both residents and staff is critical for outbreak control in LTCs. Individuals with high contact activity were consistently associated with a greater probability of initiating large outbreaks, highlighting the importance of accounting for contact heterogeneity when designing surveillance and infection-control measures. More broadly, this study demonstrates the importance of accounting for contact-network heterogeneity when designing infection prevention and control measures in LTC settings, and highlights the value of integrating empirical contact data with transmission modelling to inform evidence-based outbreak preparedness, targeted surveillance, and infection-control strategies in long-term care facilities.

## 1. Introduction

During the COVID-19 pandemic, long-term care facilities (LTCs) worldwide were heavily hit, witnessing some of the highest reported incidence and mortality rates among their residents[1]. In the UK, an early analysis[2] showed that around 6 in 10 LTCs experienced at least one excess death up to August 2020; while a later comprehensive study of 4.3 million adults estimated that during the first wave of the pandemic, individuals aged 65 years or older who resided in LTCs in England experienced an estimated 18-fold higher mortality rate in comparison to those living in private homes[3]. Globally, it’s estimated that people living in LTCs accounted for around 30% of total COVID-19 deaths (range: 9-64%) based on datasets from 25 countries[4], though this is likely to be an underestimate given the limited testing capacity during infection waves. On the other hand, working with patients in healthcare settings (both hospitals and LTCs) is also known to significantly impact the occupational health of caregivers during the COVID-19[5, 6].

This disproportionate burden in LTCs is likely caused by the inherent high vulnerability of dense concentrations of frail and/or elderly residents and frequent individual interactions for caring purposes. Unlike in the general community, many of these contacts are essential for delivering care and therefore cannot be substantially reduced through social distancing or other contact-reduction measures. To inform public health decision-making in this congregate population, mathematical models have been employed to assist in understanding the epidemiological processes throughout the pandemic[7]. These models helped answer epidemiological questions in (near) real-time, but at the expense of oversimplifying transmission dynamics and population behaviours due to inadequate data, particularly during earlier stages of the pandemic[7]. One key feature many models failed to incorporate was individual contact behaviours. A recent study[7] reviewed models used globally for informing SARS-CoV-2 infection control within LTCs and found 72% (18/25) of such models claimed they considered individual contact behaviours by using agent-based or network model frameworks. However, the majority of these (12 out of 18) assumed contact patterns arbitrarily: either using estimates from pre-pandemic literature or expert opinions. This highlights a critical gap in our understanding of contact networks within LTCs, given that the uncertainty around contact patterns could significantly impact outbreak outcomes and the corresponding effectiveness of different interventions[8].

Unlike the mainly unstructured contact patterns from the general population, the contact patterns in healthcare settings (both LTCs and hospitals) are relatively structured, well-defined due to frequent care provision interactions, and could be captured via different data collection methods. Although previous studies have attempted to use wearable sensors[9–11], self-reported diaries[12], or electronic medical records[13] to monitor contact dynamics within care settings, these studies have been confined largely to hospital environments or have captured data for only small population samples and brief durations (less than 5 days). Therefore, there is still a notable scarcity of observational data on contact patterns, specifically within LTC settings.

In this study, we aim to explore how incorporating detailed contact network structures into transmission models would deepen our understanding of infectious disease outbreaks in LTCs and could inform outbreak management. We first present a detailed analysis of real-world contact patterns from care delivery, as recorded in electronic health records from a UK long-term care (LTC) unit. By utilizing this empirical contact data, we then constructed a stochastic network-based transmission model, which was further employed to simulate infections that resemble respiratory pathogens or other infections spread through close contact and to examine the effectiveness of symptom-based interventions for outbreak control. The results of this study have implications not only for the management of COVID-19 outbreaks in LTCs, but also for the development of effective strategies for pandemic preparedness and response that account for the complex and heterogeneous nature of contact networks in different populations.

## 2. Methods

### (a) Data

We studied one unit of an LTC located in London, UK, from April to May 2020 (62 consecutive days), during the first wave of the COVID-19 pandemic, when no vaccination was available and infection control measures, including visitor restrictions and social distancing policies, were in place. The unit was physically separated from the rest of the home, and the residents were not in contact with residents in other units. In March 2020, the staff were also set up in a cohort and only cared for residents in the unit. During that time, 32 nurses (25 permanent staff and 7 agency staff, i.e. temporary nurses employed through external staffing agencies) worked with 18 care home residents in a single unit of the care home. Each member of staff carried an electronic device to log interactions with care home residents to provide service and care. In total, there are 28,282 service events recorded in the dataset. The contact information was collected through the secondary service records where staff registered their interaction type and time with residents on digital devices when providing service. Typical service interactions include providing meals, medication, comfort rounds, and activities. This studied LTC unit is designed for the elderly population and is of the median size of similar UK facilities[14]. The data were originally collected for care management purposes and not specifically for research. They were obtained from a care home database after anonymisation under strict data protection protocols agreed between the University of Exeter and the care home provider. The data were shared with the University of Oxford and the University of Bristol in an anonymised form, in accordance with that data sharing agreement. The ethics of the use of these data for these purposes was agreed by Public Health England with the UK government SPI-M(O)/SAGE committees.

### (b) Empirical contact network construction

The electronic service record documented details of which staff provided what type of service to which resident and at what time. As staff would provide multiple services during one visit to the resident (i.e. to provide drugs while offering lunch), we thus defined a staff and a resident to be in ‘contact’ when a service is recorded, and merged service records within 10-minute intervals into one unique ‘contact’. This contact information was further used to construct an hourly temporal contact network in which nodes represent individuals and edges between nodes represent a contact. The duration of contacts was assumed to follow one unit of time step (one hour) for computation. We also constructed two weighted aggregated networks (for the whole studied duration and for the first 3 days, respectively) where the edge weight is proportional to the total number of contacts between the corresponding 2 individuals. Figure S1 shows the details regarding data processing. Chi-squared tests were used to assess whether there were significant differences in the distribution of service types between staff type. We noted that since there were no staff-to-staff and resident-to-resident services provided, our contact data contains only necessary contacts for care purposes, underestimating the contact rates within this LTC.

For the constructed hourly and two aggregated networks, we first characterize the network properties by computing several classical global graph metrics, including *Total number of nodes, Total number of edges, Edge density, Sum of weights, Diameter*, and *Average shortest path length*. We also analyse the importance of individuals in terms of ‘how connected’ or ‘how central’ a node is within a network by computing the local network metrics. Specifically, we include *degree, betweenness, eigenvector centrality*, and *strength*. Each metric provides a different measure of the centrality of a node, and the detailed calculations and interpretations of these metrics can be found in the Supplementary information. Nodes were ranked based on their centrality values for identifying key influencers and the Kruskal-Wallis test or the Mann-Whitney U test was applied to explore if significant difference exists among different individual types (residents, permanent staff, and agency staff). The network analysis was conducted in R (version 4.3.0) using *igraph* package[15].

### (c) Epidemic simulations

We built a stochastic network model, where the infection process goes through 5 infection stages: Susceptible (S), Exposed (E), Symptomatically infected (I), Asymptomatically infected (A), and Removed (R) (Figure S2). We parameterised life history parts of the model based on SARS-CoV-2 transmission and disease progression estimates found in the literature[16]. The model assumes a confined environment with no birth or natural or disease-induced death, and individuals do not alter their behaviour after becoming infected, so the edges do not rewire in response to disease progression. Given visiting restrictions during outbreaks, we did not include visitors in the network.

Although there have been many estimates of the reproduction number of SARS-CoV-2, very few of these are for this type of closed setting [17, 18]. Therefore, we varied the probability of transmission per contact occurring in a one hour period (β) and investigated the effect of changes in this parameter on the outbreak dynamics.

Simulations were run on the hourly empirical temporal contact network described above with a single initial infectious individual (who may or may not seed the outbreak) selected at time step 1. The empirical network changed at every time step, as reflected in the data, with new edge lists (contact) of susceptible and infectious individuals generated. The simulations then unfold stochastically with transmission events determined through a random binomial draw of a function of transmission probability per contact (β). Both the infectious period and incubation period were randomly drawn from gamma distributions once the transmission event occurred. Our model codes are based on the R package *EpiModel (v2*.*3*.*2)*[19], which is a flexible platform allowing user-defined infectious disease dynamics and network properties (see Supplementary). 1000 simulations of the model were run for each distinct scenario (differed by starting points and key parameters).

We explored two types of scenarios: (a) consider different routes of importation to the LTC by varying the infectious seed, and (b) consider different transmission probabilities that represent different social distancing intensities by varying the transmission probability per contact (β). An outbreak was defined as at least one other individual infected by the infectious seed, and we plotted the probability of an outbreak across the 1000 simulations. We computed the following outcome measures for each simulation and reported the distributions of these outcomes over 1000 simulations: infection incidence, outbreak final size (cumulative number of cases in each simulation), outbreak full duration, peak duration and underlying incidence when observing 2 symptomatic cases. We also compute average secondary infections per day to understand super-spreading events. For scenario (a), we excluded the agency staff as their data points may refer to multiple people.

## 3. Results

### (a) Contact patterns

#### (i) Repeated daily service provision patterns

There are overall 28,282 service events between 18 residents, 25 permanent staff and 7 agency staff over 62 days recorded in the dataset. The staff delivered care services inside the studied LTC on a daily basis, and the corresponding repeated service provision patterns for typical services are outlined in Table S2. For a typical day, staff would start their 12-hour day shift at around 7-8 AM, provide 3 meals and medication for all residents at the scheduled time, and deliver other care services (comfort rounds, activities, special checks, etc.) based on residents’ distinct needs. The staff members undergo a shift change at around 7-8 PM to commence the night shift, during which the primary activity involves conducting checks every two hours throughout the night. These services were jointly provided by the agency staff and permanent staff, depending on the composition of staff members. It’s observed that the service types provided by agency staff were significantly different from those provided by permanent staff (Chi-squared test, p<=0.01), as shown by contact matrices illustrating interactions per hour between agency staff, permanent staff, and residents, broken down by different services (Figure S3 and Figure S4), and overall the permanent staff recorded a noticeable higher number of ‘activity’ and ‘medication’ services compared to the agency staff.

#### (ii) Contact heterogeneity across time and individual

Due to the varying health statuses of the residents and shift patterns, the interactions between residents and staff members resulting from service provision exhibit considerable variation. A particular service could be provided by either a single staff member or multiple staff members collectively (i.e. a single staff member may serve lunch to multiple residents, while checks may be conducted by multiple staff members). Figure 1a and Figure 1b highlight the observed heterogeneity across individuals and time by displaying service counts for the same person on 2 different days and 3 hourly contact networks. Figure 1a also illustrates the shift patterns for staff’s service delivery, where service counts were relatively higher during day shifts and service types were relatively limited during night shifts. Overall, each resident had an average of 21 (IQR:17-25) distinct contacts with staff and encountered 5 (IQR:4-6) unique staff members per day. In extreme cases, one resident could meet up to 11 different staff over one single day and receive 56 services at various hours.

**Figure 1.**
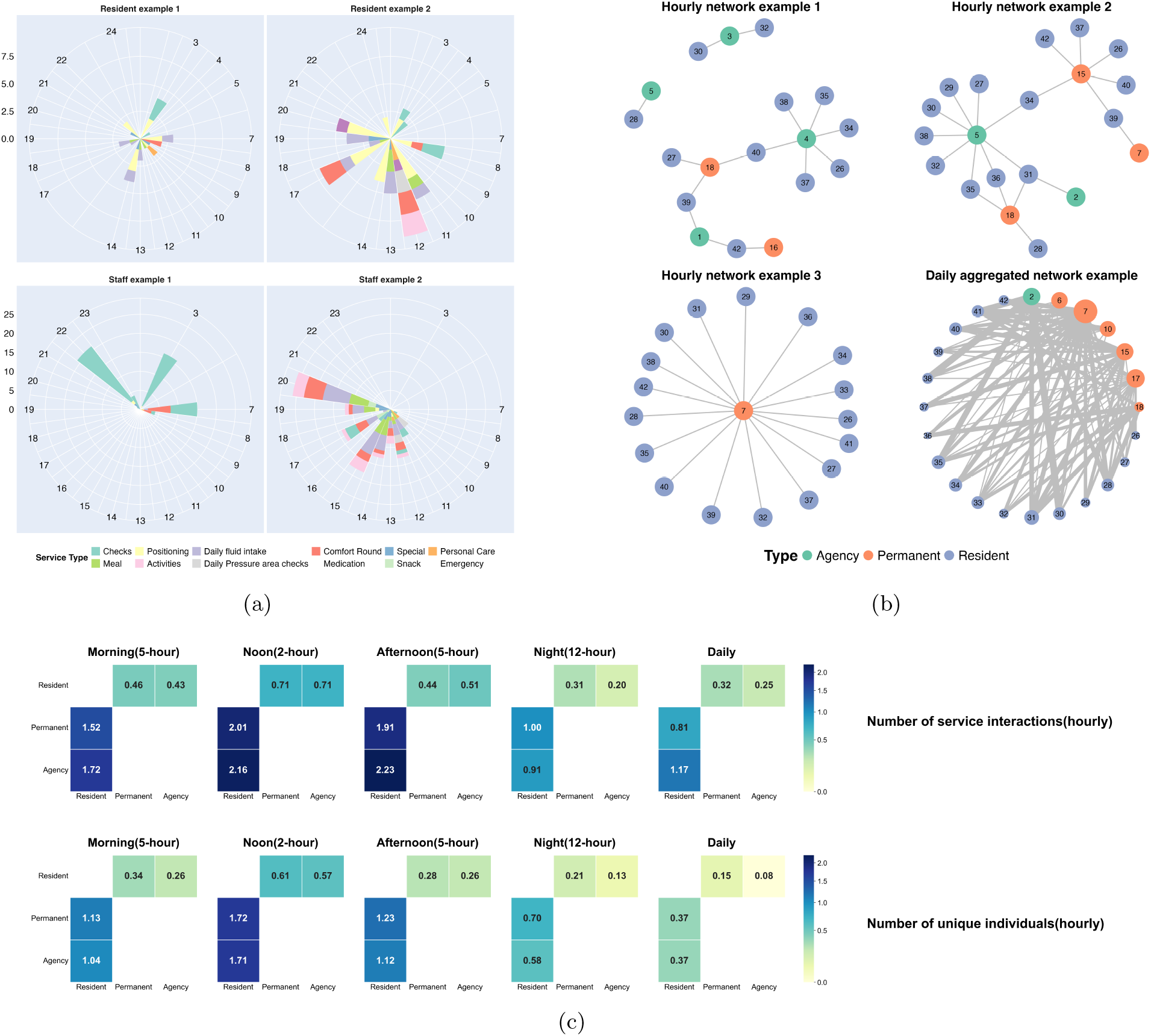
Contact heterogeneity across time and individuals. **(a)** Distribution of services received by example individuals, broken down by service type (colour) and measured hourly (radial bars) across different days. **(b)** Three selected hourly contact network snapshots and one daily aggregated network. Nodes represent individuals and are coloured by individual type, while edges indicate contacts. In the daily aggregated network (bottom right), node size represents each individual’s total interaction frequency, and edge thickness represents the interaction frequency between each pair of individuals. **(c)** Hourly contact matrices for four time periods and the full day. Contact is defined by the number of service interactions in the top row and by the number of unique individuals contacted in the bottom row.

As we observed high heterogeneity among individuals’ contact patterns, we thus used the hourly contact networks in simulations and reported the hourly contact matrices after data cleaning for the different time periods in Figure 1c. The time periods are classified into Morning(7-12 AM), Noon(12-2 PM), Afternoon(2-7 PM) and Night(7 PM-7 AM) based on observed patterns of contact frequency discussed in Section (a)i. The ‘contact’ are counted and visualized by either the number of service interactions one had or the number of unique individuals one met, given the nature that one staff could deliver care to various individuals and one service could be provided by multiple staff. As the utilized service records didn’t collect information on staff-to-staff and resident-to-resident interactions, the corresponding entries in the contact matrices remain empty. Both types of the contact matrices are not symmetric.

### (b) Network properties

After data cleaning, there were 13,101 unique contacts between 43 individuals over 62 days. In Figure 1b, we visualized the various networks (the hourly and daily) generated based on these data and documented key network metrics for the weighted daily networks in Table S3. Overall, the aggregated network over the whole study period is more extensive, more interconnected, and with stronger interactions between nodes compared to the daily networks. However, both the aggregated and daily networks showed limited clustering and no large connected components. This likely reflects the bipartite nature of the recorded care-delivery contacts, which did not include staff–staff or resident–resident interactions.

In both networks, the degree distributions were right-skewed and highly heterogeneous, which aligns with the observed heterogeneity in service counts. In terms of node importance, Nodes were ranked by these centrality measures, and notably, the top three highest-ranked nodes, and seven out of the top ten, were permanent staff members, suggesting their potential role as key influencers in this community. Further statistical analysis showed significant differences in the median ranks across node types (Kruskal-Wallis test, p*<*0.05). Specifically, resident nodes ranked significantly higher when ranking based on degree centrality or strength compared to both agency and permanent staff nodes (Mann-Whitney U Test, p*<*0.05). This implies that if residents become infected, they may be more influential in rapidly spreading the infection within the contact network as these two metrics measuring the direct connectivity to other nodes. In contrast, nodes of permanent staff ranked significantly lower in betweenness centrality than nodes of residents and agency staff (Mann-Whitney U Test, p*<*0.05), suggesting they are less likely to serve as bridges connecting disparate groups. Additionally, agency staff nodes ranked significantly lower in eigenvector centrality than residents and permanent staff nodes (Mann-Whitney U Test, p*<*0.05), indicating they are less connected to other highly connected, influential nodes.

### (c) Network model simulation results

#### (i) Seeding outbreaks

Upon developing a transmission model integrating the temporal contact networks, we investigated the potential infection transmission dynamics by evaluating two distinct scenarios: (a) examining different routes of viral importation to this LTC, and (b) assessing the influence of variable transmission probabilities, which could reflect differing levels of virus infectivity and/or social distancing measures. Overall, our simulations show that whether an outbreak will happen depends not only on the transmission parameter but also on the characteristics (type and level of contacts) of the initial seeding individual (Figure 2(a)). As expected, the probability of a large outbreak (case number > 10) generally increases when the probability of transmission per contact increases. For example, when the transmission probability is 0.05, 27.4% of simulations(aggregating all seeding scenarios) lead to a large outbreak, whereas when the transmission probability increases to 0.5, 63.5% of simulations result in a large outbreak. Outbreaks are very unlikely when transmission probability is less than 0.01 (Probability of a large outbreak < 0.5%) but caps at around 65% even when transmission probability per contact is larger than 0.5(0.5:63.5%;0.75:65.2%; 0.99:66.1%). Notably, while outbreaks can be be triggered by any individual, simulations seeded from individuals with higher contact activity generally showed a greater probability of generating large outbreaks. Figure 2(a) illustrates this pattern using four example seed individuals. For example, when the transmission probability was 0.99, the probabilities of a large outbreak were 12.0%, 68%, 90%, and 93% for the low-degree resident, high-degree resident, low-degree staff member, and high-degree staff member, respectively. Even at a more moderate transmission probability of 10%, introduction of infection through the high-degree resident and high-degree staff member shown in Figure 2(a) resulted in a greater than 75% probability of a large outbreak.

**Figure 2.**
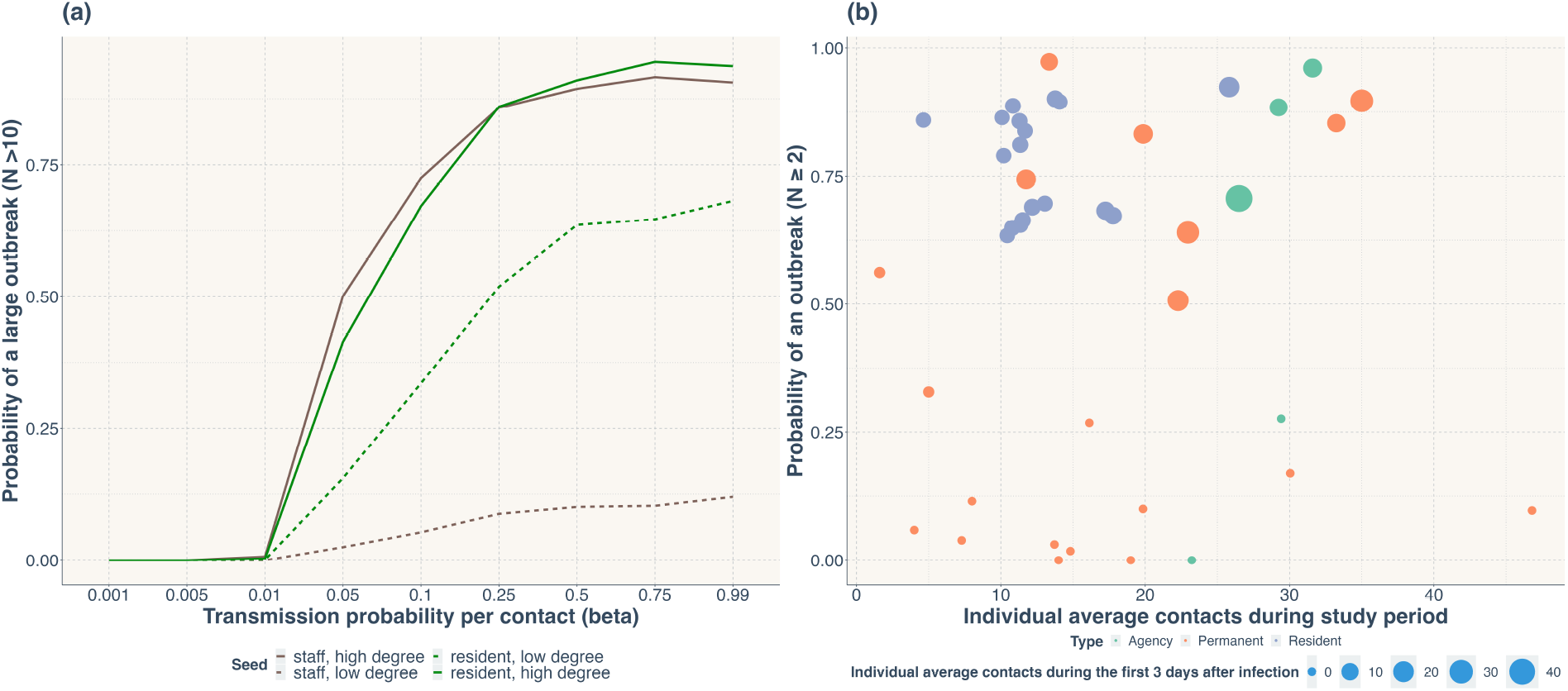
Probability of an outbreak. **(a)** probability of a large outbreak (N>10) across different transmission probabilities per contact (β), for four example seed individuals: one high-degree resident (green, solid line), one low-degree resident (green, dashed line), one high-degree staff member (brown, solid line), and one low-degree staff member (brown, dashed line). **(b)** probability of an outbreak (N ≥ 2) for a fixed transmission probability per contact (β= 0.5), for simulations seeded on individuals with different degrees in the hourly temporal network, with the number of contacts of the seed individual in the first 3 days of the outbreaks indicated by the size of the bubble.

Our simulations also show that an individual’s average contacts during the first 3 days after infection could better predict the probability of an outbreak compared to the average contacts during the study period, as shown in Figure 2(b). Larger bubbles, denoting more contacts in the first three days after infection, generally align with higher outbreak probabilities, suggesting that early contact frequency may play a more significant role in spreading the infection. However, we didn’t observe a significant association between node importance (characterized by metrics such as *degree, betweenness, eigenvector centrality*, and *strength*) and the probability of outbreak occurrence during the simulations. This suggests that these metrics, which describe the positioning and connectivity of nodes within the network, do not alone predict whether an outbreak will occur.

#### (ii) Variable outbreak dynamics

The heterogeneity of contact patterns (discussed in (a)ii) further drives the heterogeneous outbreak dynamics. Figure 3 illustrates 3 simulation outcomes (epidemic curves, final size and number of secondary infections) in scenarios with transmission probability of 0.01, 0.05 and 0.25, while all simulation results can be found in Supplementary figures. As the transmission probability per contact increases, overall bigger and faster epidemics are observed (Figure 3(a) and (b)). The final outbreak size of simulations is highly scattered when the transmission probability is relatively high(0.05), as shown in the middle facet of Figure 3(b), and this variability is also pronounced when seeding infections on low-degree residents with high transmission probability(0.25). For these 2 βscenarios, the average final sizes are 7.36 and 21.28, with standard deviations of 9.79 and 17.12, respectively. Additionally, the size of the outbreaks seems more dependent on contact level compared to the type of the individuals(staff or resident). The narrowest final size distribution was observed if seeding from low-degree residents, pointing to a more limited potential for disease spread among this group. Finally, figure 3(c) shows that individuals who get infected earlier generally have higher individual reproduction numbers, but super-spreading events could also happen at a later stage.

**Figure 3.**
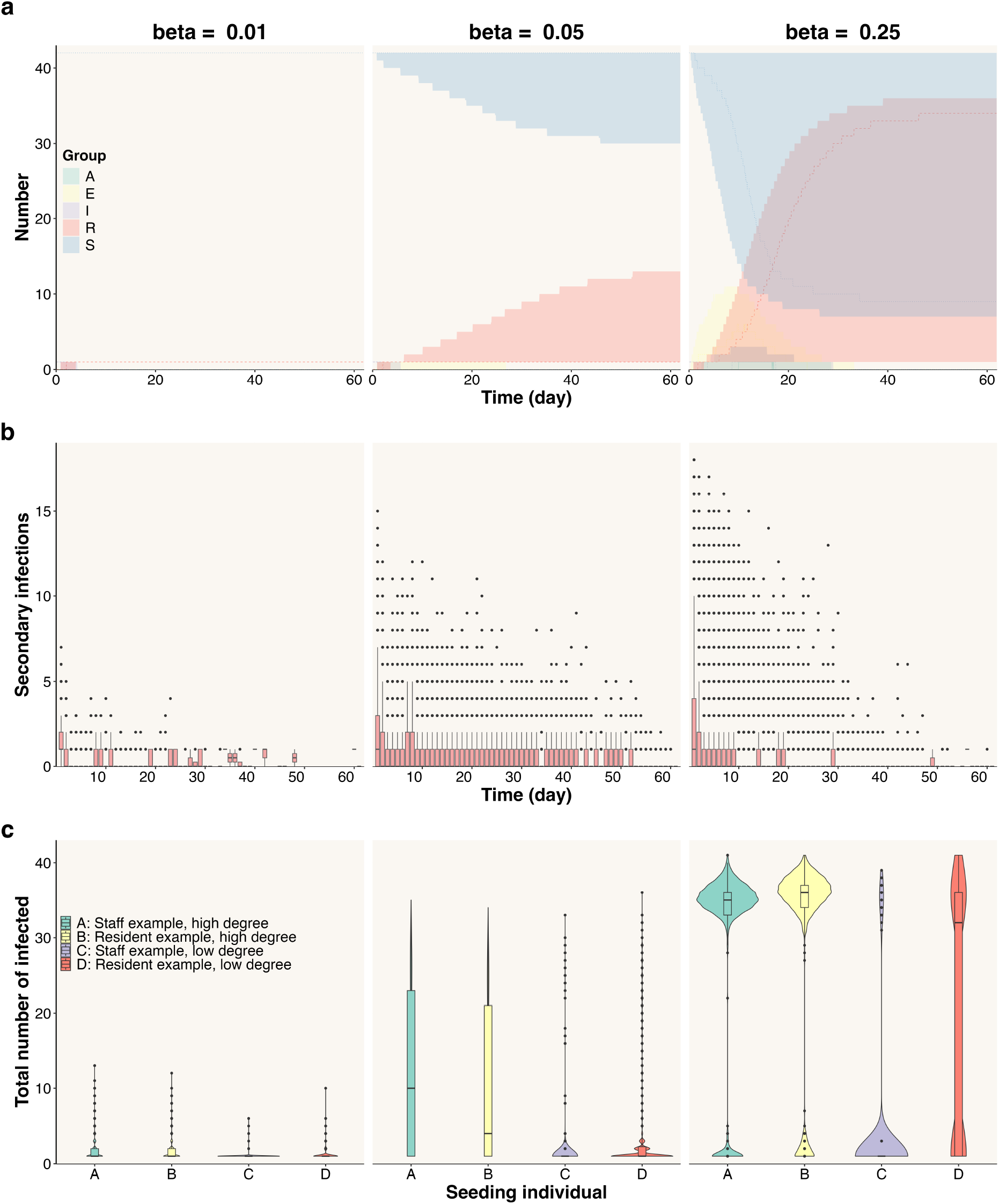
Outbreak outcomes for different transmission probabilities per contact (*β*). **(a)** Temporal epidemic dynamics for the five epidemiological compartments (A, E, I, R, and S). For each transmission probability, data were pooled across the four selected seed individuals, with 1,000 simulations per seed. Lines show the median number of individuals in each compartment at each time point, and shaded bands show the interquartile range (25th–75th percentiles).**(b)** Distribution of the number of secondary infections caused by each infected individual, according to the day on which that individual became infected.**(c)** Distribution of the total number of individuals infected by the end of each simulation (final size). Violin plots and embedded boxplots are shown separately for four seed individuals: A, staff example with high degree; B, resident example with high degree; C, staff example with low degree; and D, resident example with low degree.

#### (iii) Symptom-based testing

Finally, we explored whether our network-based transmission model could help to optimize the use of control measures, such as symptom-based testing, to minimize outbreaks in LTCs. For this, we analysed the time of the presentation of infection symptoms for different transmission setups. As the transmission probability per contact (β) increases, the time taken to observe 2 symptomatic cases decreases and the underlying incidence increases(Figure 4). In the most severe scenarios (seeding infection from high-degree staff with β=0.99), the average number of underlying cases when observing 2 symptomatic cases is 13.12 (SD:5.63), which suggests that detecting an outbreak at this stage would likely already be too late to control the epidemic. We also found that the average time required for the presentation of 2 symptomatic cases could be as low as 1.5 days or as high as 19.4 days, which again raises concerns about the practicality of symptom-based testing as an early containment strategy. This is especially critical because delays in detecting and responding to outbreaks can lead to widespread transmission, making containment increasingly challenging. These patterns persist across a broad spectrum of transmission probabilities. Notably, when infection was seeding from low-degree staff, the increased transmission probability does not correspondingly decrease the detection time for two symptomatic cases (Figure 4). This anomaly could suggest that contact frequency holds greater sway in outbreak trajectories than mere transmission probabilities, especially where individuals with fewer contacts are less efficient in spreading the infection. This emphasizes the importance of targeting high-contact individuals in prevention and control strategy design.

**Figure 4.**
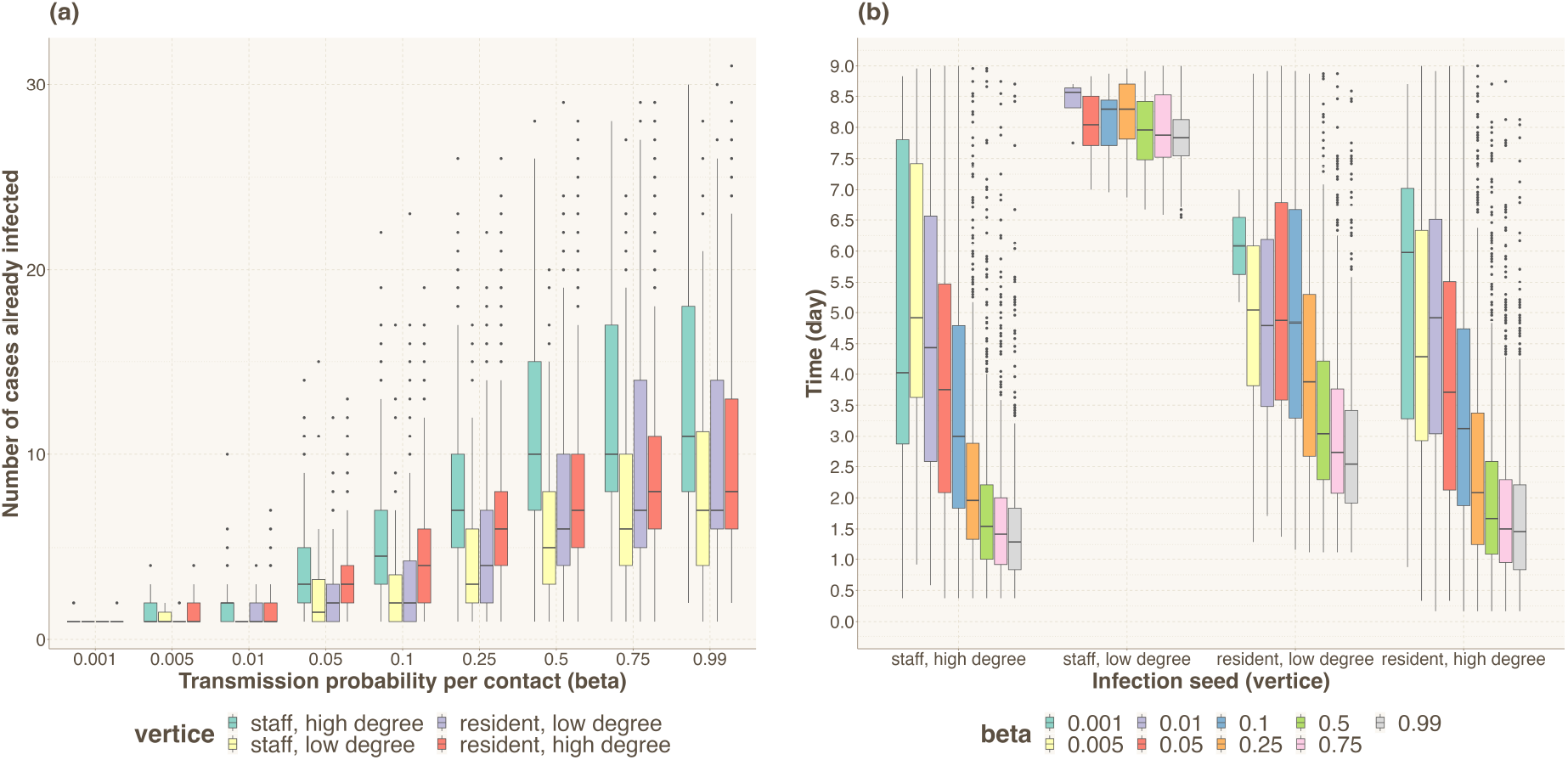
Symptom-based testing. **(a)** Incidence when observing two symptomatic cases, stratified by individual type (color), across different transmission probabilities (β) **(b)** Time to observe two symptomatic cases in simulations resulting in at least two symptomatic cases, categorized by individual type, plotted against different transmission probabilities (β, color)

## 4. Discussion

While the COVID-19 pandemic disproportionately affected residents in LTCs, contact patterns and infection dynamics within these settings remain poorly understood, limiting the utility of current transmission models. Here, we characterized a highly heterogeneous contact network observed in a UK care home during the first two months of the COVID-19 pandemic and analysed care provision patterns as influenced by staff shifts and residents’ care needs. This extended observation period enabled us to characterise contact heterogeneity more extensively than has been possible in previous studies. We also presented a stochastic transmission model parameterized using the observed contact networks and showed that infection transmission dynamics can be highly variable once contact heterogeneity is incorporated. Although the contact data were collected during the COVID-19 pandemic and SARS-CoV-2 serves as a motivating example throughout the manuscript, the transmission model was not calibrated to represent COVID-19 specifically. Instead, transmission probabilities were varied to explore how observed contact-network structures influence transmission dynamics across a range of plausible outbreak scenarios. To the best of our knowledge, this study provides one of the most detailed high-resolution analyses of contact patterns relevant to respiratory and close-contact infections in LTC settings. Therefore, the insights gained from our modelling have implications not only for understanding SARS-CoV-2 transmission in LTCs but also for informing preparedness and infection control strategies for other respiratory pathogens in similar settings.

Staff and residents in our studied LTC recorded contact rates higher than the general UK community over the same time period[20], and similar to rates reported in caring settings elsewhere[9, 11, 12], despite slight differences in the definitions of contacts. The raw data revealed high-frequency daily interactions, with residents receiving an average of 21.64 contacts (IQR: 17.00–25.00) from approximately five unique staff members daily. The observed repeated daily service provision patterns aligned with the shifts and care needs of this community. However, the degree of contact heterogeneity observed across different times and among individuals demonstrates the complex contact dynamics within LTC environments, highlighting the need for infection control measures that account for this variability. We also noted that, on average, the contact level of staff members was higher compared to residents, and the types of services provided were not significantly correlated with the categories of staff involved. This indicates that, in the studied LTC, service provision was organised based on need and convenience rather than following specific staff-role requirements. To facilitate future research, we have provided detailed service summaries and contact matrices in the Supplementary Information, allowing others to incorporate empirically observed care-delivery patterns into alternative modelling frameworks.

Interactions and contacts between individuals can be conveniently represented within a network framework, where nodes represent individuals and links correspond to observed contacts. Analysis of the aggregated contact networks showed that the network structure was relatively stable from day to day, with a small number of highly connected individuals, typically permanent staff members, accounting for a disproportionate share of contacts. On average, approximately 60% of participants were active on any given day, while only around 27% of all possible connections were utilised. The average path length suggests that an individual would typically need to pass through only a few intermediate individuals to reach any other person within the network. At the same time, considerable heterogeneity was observed in individual connectivity, indicating unequal opportunities for transmission between different individuals and potentially contributing to the variability in outbreak outcomes observed in our simulations. Temporal network analysis further showed that contact structures varied throughout the day, largely reflecting staff schedules and changing care demands. Consequently, the observed network structure depends strongly on the temporal resolution at which contacts are analysed, and aggregating contacts over longer periods may obscure important transmission-relevant dynamics. It should be noted that the recorded contact network reflects care-delivery interactions between staff and residents only, and therefore has a bipartite-like structure with no recorded staff-to-staff or resident-to-resident contacts. This should be taken into account when interpreting global network properties such as clustering and connectedness. Together, these findings highlight the importance of considering both contact heterogeneity and temporal variation when modelling infection transmission in LTC settings.

Using our simulation model, we investigated the potential transmission outcomes by varying the initial source of infection (infection seed) and the transmission probability per contact. The simulations revealed that outbreak dynamics were highly heterogeneous and difficult to predict. Introduction from either staff or residents could lead to large outbreaks, although the resulting outbreak size, timing, and duration varied considerably across different scenarios. The duration of the simulated epidemic ranged from as brief as 2.5 weeks to as prolonged as 8.5 weeks. A particularly important finding was that individuals with higher contact activity were more likely to generate large outbreaks following infection introduction, reflecting the heterogeneous connectivity observed in the contact network. This suggests that outbreak outcomes in LTC settings are influenced not only by pathogen characteristics but also by the structure of the underlying contact network and the position occupied by the infection seed within that network.

At lower transmission probabilities, outbreaks were much more likely to die out by chance before extensive spread occurred. The slower transmission dynamics also provide more opportunities for intervention, provided that infection is not introduced into highly connected parts of the network at an early stage. We also showed that symptom-based testing interventions are unlikely to be effective at detecting outbreaks early, as the time taken to observe two symptomatic cases could be substantial while the underlying incidence may already be high. This is consistent with experiences reported from LTC outbreaks during the COVID-19 pandemic[21, 22], where silent introduction and pre-symptomatic transmission contributed to large outbreaks despite symptom-based surveillance and contact restrictions. Our findings are broadly consistent with the existing literature, which emphasises the importance of early detection and the use of multiple complementary interventions to control infection in LTC settings[7]. However, the observed heterogeneity in contact behaviour suggests that transmission risk may not be distributed evenly across individuals. While we did not explicitly evaluate alternative testing strategies, our results indicate that individuals with consistently high levels of contact may contribute disproportionately to outbreak risk following infection introduction. This may help explain why previous modelling studies have reached different conclusions regarding whether staff or residents should be prioritised for routine screening, as such recommendations depend strongly on assumptions regarding contact behaviour. In settings where surveillance resources are limited, approaches that account for individual contact activity may warrant further investigation.

One key parameter in our simulations is the probability of transmission per contact, which encapsulates factors such as individual susceptibility, infectiousness, contact intensity, and the effectiveness of infection control measures. This remains a key unknown for LTC settings, and we assumed it to be uniform across all contacts. The transmission probabilities considered in this study were not chosen to represent any specific pathogen; instead, they were varied to explore how differences in transmission potential interact with the observed contact-network structure to influence outbreak dynamics. For highly heterogeneous contact networks, there is no simple one-to-one mapping between the per-contact transmission probability and the basic reproduction number (*R*_0_), as transmission outcomes depend not only on the transmission probability itself but also on the structure of the contact network and the position of infected individuals within it. Further research could explore whether incorporating service-specific transmission risks alters outbreak dynamics and intervention effectiveness.

Existing literature[7] largely agrees that a combination of interventions, with a particular emphasis on early detection, is essential for controlling infection within LTCs, and our data-driven network model supports this public health implication. We showed that symptom-based testing alone is unlikely to detect outbreaks at an early stage, as the time taken to observe two symptomatic cases could be long while the underlying incidence may already be high. However, while some previous modelling studies[23] have suggested that staff are the primary route of viral introduction, our results indicate that the degree of contact may be more important than individual type alone. Both staff and residents were capable of initiating large outbreaks, but individuals with higher contact levels were consistently associated with a greater probability of generating large outbreaks following infection introduction. This may help explain why previous modelling studies have reached different conclusions regarding whether staff or residents should be prioritised for routine screening, as such conclusions depend heavily on assumptions regarding contact behaviour. Should surveillance or testing resources be limited, our findings suggest that prioritising highly connected staff members may be a reasonable approach, as residents frequently interact with them and infection introduction through highly connected individuals was associated with a greater probability of large outbreaks. However, evaluating the effectiveness of such targeted strategies would require further modelling beyond the scope of the present study. The structured nature of the observed contact network may also have implications for cohorting strategies. Contacts were concentrated within repeated care-delivery patterns rather than being distributed uniformly across the facility, suggesting that maintaining consistent staff-resident groupings could help limit transmission opportunities between otherwise weakly connected parts of the network. Although cohorting interventions were not explicitly evaluated in this study, the observed network structure supports the rationale underlying such approaches.

This study has several limitations in terms of model assumptions and available data. First, the study was conducted within a cohorted LTC unit where residents were organised according to care needs and staffing arrangements. Contact structures may therefore differ from those observed in facilities with different layouts, staffing models, or resident populations. Secondly, there was no staff-to-staff or resident-to-resident contact information, as our contacts were based on service provision records; we also did not capture potential contacts resulting from chats or casual meetings. This likely led to an underestimation of the overall contact level within the LTC. However, the implications for infection control remain valid, as this underestimation effectively positions our model as a best-case scenario. Furthermore, agency staff were excluded from the transmission simulations because their contact patterns could not be consistently reconstructed from the available records. As agency staff may connect otherwise separated groups within and between facilities, excluding them may have resulted in an underestimation of transmission opportunities. Although we argue that the studied LTC is representative of a typical English care home, the sample size remains relatively small, and variability in individual reporting behaviour could potentially influence the observed differences in contact patterns. Whether our findings can be generalised to other settings therefore remains uncertain, given that the simulation model was tailored to the contact network of a particular LTC unit. However, we have included the repeatedly observed service descriptions and contact matrices, stratified by service type, in the Supplementary Information to enable readers to construct hypothetical facilities of interest. Finally, we did not account for visitors or consider that the likelihood of acquiring infection from the community may vary between different types of individuals. We also did not calibrate the transmission model using infection data, as no COVID-19 outbreaks occurred in the studied LTC during the observation period.

In conclusion, our study employs high temporal resolution observational data to reveal that contact networks within an English LTC are highly heterogeneous and dynamic. These variations in network structure have the potential to influence disease dynamics and individual contributions to outbreaks. Our modelling results show that, once infection is introduced, outbreak occurrence, timing, size, and the likelihood of super-spreading events can vary substantially, making transmission dynamics difficult to predict. Preventing infection introduction through both staff and residents therefore remains a key component of infection control in LTC settings. More broadly, our findings demonstrate the importance of accounting for contact-network heterogeneity when modelling infection transmission and designing outbreak prevention and control strategies in long-term care facilities.

## Data Availability

All data produced in the present study are available upon reasonable request to the authors

## Ethics

The data were originally collected for care management purposes and not specifically for research. They were obtained from a care home database after anonymisation under strict data protection protocols agreed between the University of Exeter and the care home provider. The data were shared with the University of Oxford and the University of Bristol in an anonymised form, in accordance with that data sharing agreement. The ethics of the use of these data for these purposes was agreed by Public Health England with the UK government SPI-M(O)/SAGE committees.

## Data Accessibility

The code used for analysis is available at (https://github.com/Pie923/carehome_network_model_public). Further details of our methods and results can be found in electronic supplementary material.

## Authors’ Contributions

Conceptualisation: LP, LD, TDH Formal Analysis: LP

Funding acquisition: ELD, LD, TDH

Investigation: All authors

Methodology: All authors

Software: LP

Validation: All authors

Visualization: LP

Writing - original draft: LP

Writing - review & editing: All authors

## Acknowledgements

The authors would like to thank the EpiModel team for developing the modelling framework used in this study and for their helpful support throughout this work. We are also grateful to the DPhil thesis examiners, Professor Lisa White and Professor Graham F. Medley, for their constructive comments and suggestions on an earlier version of this manuscript.

## Disclaimer

Generative artificial intelligence (ChatGPT, OpenAI) was used to assist with language editing and code organisation during the preparation of this manuscript. All scientific content, analyses, interpretations, and conclusions were developed and verified by the authors.

