## appendix for "Empirical contact networks reveal heterogeneous outbreak risks in a UK long-term care facility: a modelling study": Empirical_contact_networks_reveal_heterogeneous_outbreak_risks_in_a_UK_long_term_care_facility__a_modelling_study_app.pdf

---

### S1. Method: analysing network structure

We analysed the constructed hourly and 2 aggregated networks by computing below global graph metrics and local node metrics. While we outline here the essential computational details for understanding, a more detailed description and interpretation of these metrics can be found in Newman's textbook[1].

#### *S1.1. Global Network Metrics*

- **Total Number of Nodes** ($N$): Represents the count of unique entities(individual) in the network. It gives a basic size of the network.

[*]Corresponding authors

[1]Joint last authors

- **Total Number of Edges** ($E$): The number of connections between nodes. Here we define a connection if a contact was recorded in the dataset. It provides insight regarding 'how connected' the network is.

- **Edge Density** ($D$): Calculated as the ratio of the actual number of edges to the number of all possible edges. It describes how densely connected the network is. A high density indicates a closely connected network, while a low density suggests a sparser one. For undirected graphs, calculated as:

$$D = \frac{2E}{N(N-1)} \tag{1}$$

- **Sum of Weights** ($W_{\text{sum}}$): Refers to the total sum of all edge weights in weighted networks. It describes the total interaction volume or intensity in the network. Calculated as:

$$W_{\text{sum}} = \sum_{i,j} w_{ij} \tag{2}$$

  where $w_{ij}$ is the weight of the edge between nodes $i$ and $j$.

- **Diameter** ($\Phi$): The maximum shortest path length between any two nodes. It indicates the "size" of the network in terms of the distance between its furthest nodes, given by:

$$\Phi = \max(d_{ij}) \tag{3}$$

  where $d_{ij}$ is the shortest path between nodes $i$ and $j$.

- **Average Shortest Path Length** ($L$): The average number of steps along the shortest paths for all possible pairs of network nodes. It measures the network's efficiency in information or resource transfer. It is calculated as:

$$L = \frac{1}{\frac{1}{2}N(N-1)} \sum_{i \neq j} d_{ij} \tag{4}$$

*S1.2. Local Network Metrics*

- **Degree** ($k_i$): For a node $i$, the degree is the number of connections (edges) it has. High-degree nodes are often considered influential or hubs within the network. It is given by:

$$k_i = \sum_{j} a_{ij} \tag{5}$$

where $a_{ij}$ is an element of the adjacency matrix, indicating the presence (1) or absence (0) of an edge between nodes $i$ and $j$.

- **Betweenness Centrality** ($C_B(i)$): Measures the extent to which a node lies on paths between other nodes. It describe the node's importance via measuring a node's role in facilitating information flow between other nodes. It is defined as:

$$C_B(i) = \sum_{j \neq i \neq k} \frac{\sigma_{jk}(i)}{\sigma_{jk}} \tag{6}$$

  where $\sigma_{jk}$ is the total number of shortest paths from node $j$ to node $k$ and $\sigma_{jk}(i)$ is the number of those paths that pass through node $i$.

- **Eigenvector Centrality** ($C_E(i)$): Reflects the influence of a node based on the principle that connections to high-scoring nodes contribute more to the score of the node in question. It is calculated as:

$$C_E(i) = \frac{1}{\lambda} \sum_{j} a_{ij} x_j \tag{7}$$

  where $x_j$ is the centrality of node $j$ and $\lambda$ is a constant.

- **Strength** ($s_i$): For weighted networks, the strength of a node is the sum of the weights of its edges.It reflects the node's overall connectivity strength, considering both the number and the weight of connections. It is defined as:

$$s_i = \sum_{j} w_{ij} \tag{8}$$

  where $w_{ij}$ is the weight of the edge between nodes $i$ and $j$.

### S2. Method: simulations

The network model in this study was coded in the R using the *Epimodel*(v2.3.2)[2] software platform. This software provides a flexible framework and usually comprises two steps at each discrete time interval. First, it simulates a dynamic network adhering to specified target statistics through the use of Exponential-family Random Graph Models (ERGMs); then, it models epidemics over these dynamic networks under the assumption of independent processes by simulating epidemiological transitions. We adapted this framework by replacing the ERGMs model with the observed network with all nodes and edges observed over the 1,486 time step(as recorded in the service record). The detailed code and functions are available online.

We parameterized the disease transmission process based on literature on COVID-19. Key parameters and the responding reference can be found in the below table.

| **Parameter** | **Estimate** | **Details and reference** |
|---|---|---|
| Probability of transmission per contact, $\beta$ | [0,1] | Varied by scenario. |
| Number of contacts per time step, $c$ | Variable | Based on empirical contact network data from the observed care home setting. |
| Proportion of infections asymptomatic, $p$ | 50% | [3]. |
| Incubation period, $1/\gamma$ | 5.5 days | [3]. |
| Symptomatic infectious period, $1/\sigma_1$ | 3 days | Set lower to reflect fewer contacts during the infectious period due to symptoms. |
| Asymptomatic infectious period, $1/\sigma_2$ | 3 days | |
| Relative infectiousness of asymptomatic individuals | 1 | Assumed to be equal to symptomatic value based on similar viral load levels[?] . |
| Initial infected individual (seed case) | | Varied by scenario. |

Table S1: Model parameters and assumptions used in the transmission simulations.

### S3. Supplementary result: figures

### S4. Supplementary tables

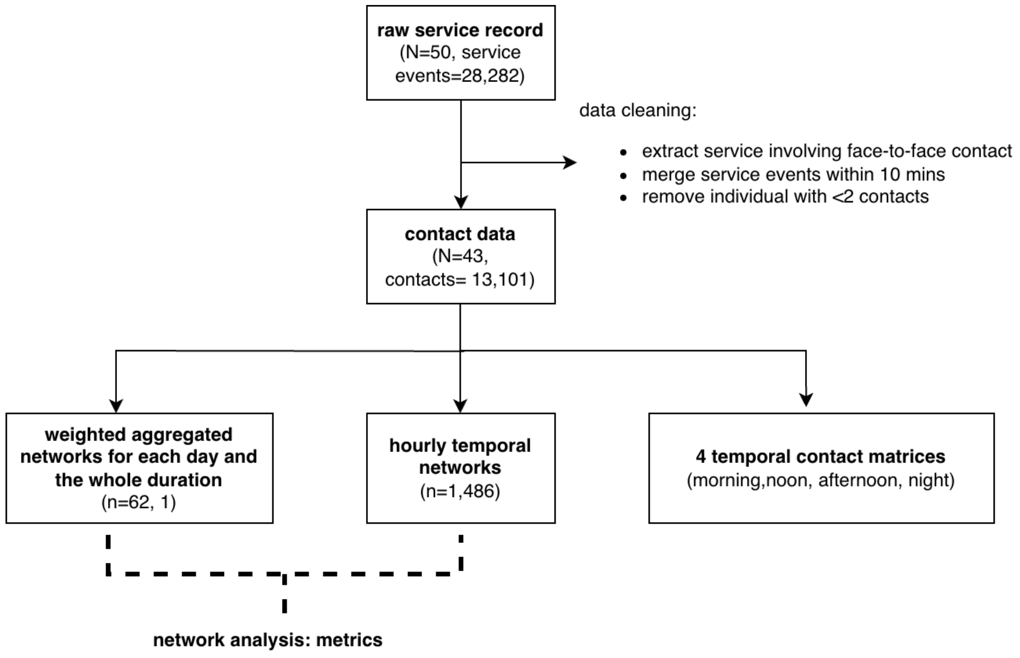


Figure S1: Data processing and network construction workflow. Raw care service records (28,282 service events involving $N = 50$ individuals) were processed to derive contact data. Data cleaning included extracting face-to-face contacts, merging service events occurring within 10 minutes, and removing individuals with fewer than two contacts, resulting in 13,101 contacts among $N = 43$ individuals. The processed data were used to construct daily and overall weighted aggregated networks, hourly temporal networks ($n = 1,486$ network snapshots), and four temporal contact matrices representing morning (7 am $\leq t <$ 12 pm), noon (12 pm $\leq t <$ 2 pm), afternoon (2 pm $\leq t <$ 7 pm), and night (7 pm $\leq t <$ 7 am the following day). These network representations were subsequently used for network analysis and transmission modelling.

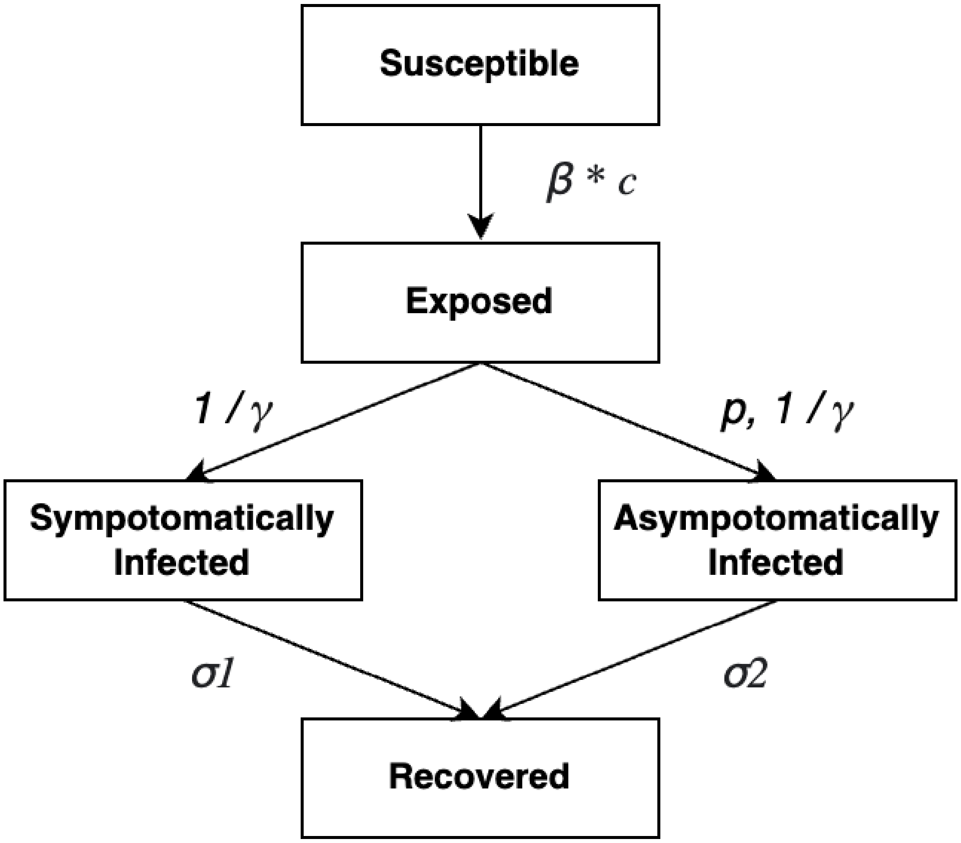


Figure S2: Structure of the stochastic transmission model. Susceptible individuals become exposed following infectious contact ($\beta c$). After a latent period ($1/\gamma$), exposed individuals develop either symptomatic or asymptomatic infection with probabilities $(1 - p)$ and $p$, respectively, before recovering at rates $\sigma_1$ and $\sigma_2$.

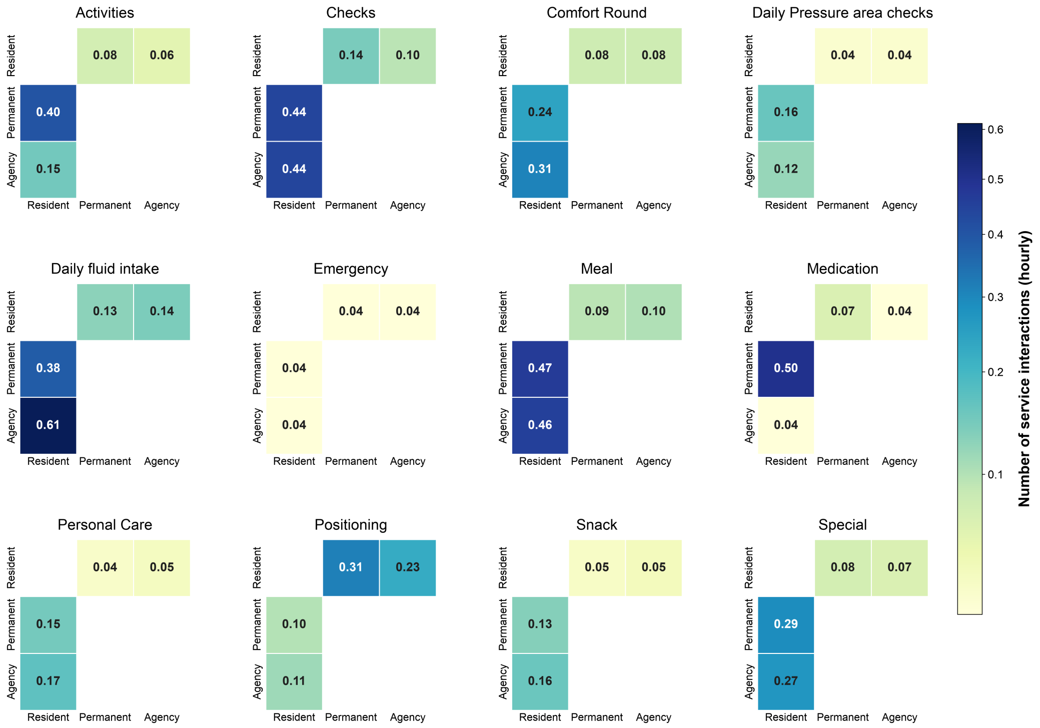


Figure S3: Service-specific hourly contact matrices using the number of service interactions as the contact definition. For each service category, matrices were constructed between residents, permanent staff, and agency staff using the processed care service records. Matrix cells represent the mean number of service interactions per hour, averaged over the observation period and normalised by time at risk. Colour intensity indicates the hourly mean, with darker blue representing more frequent service interactions.

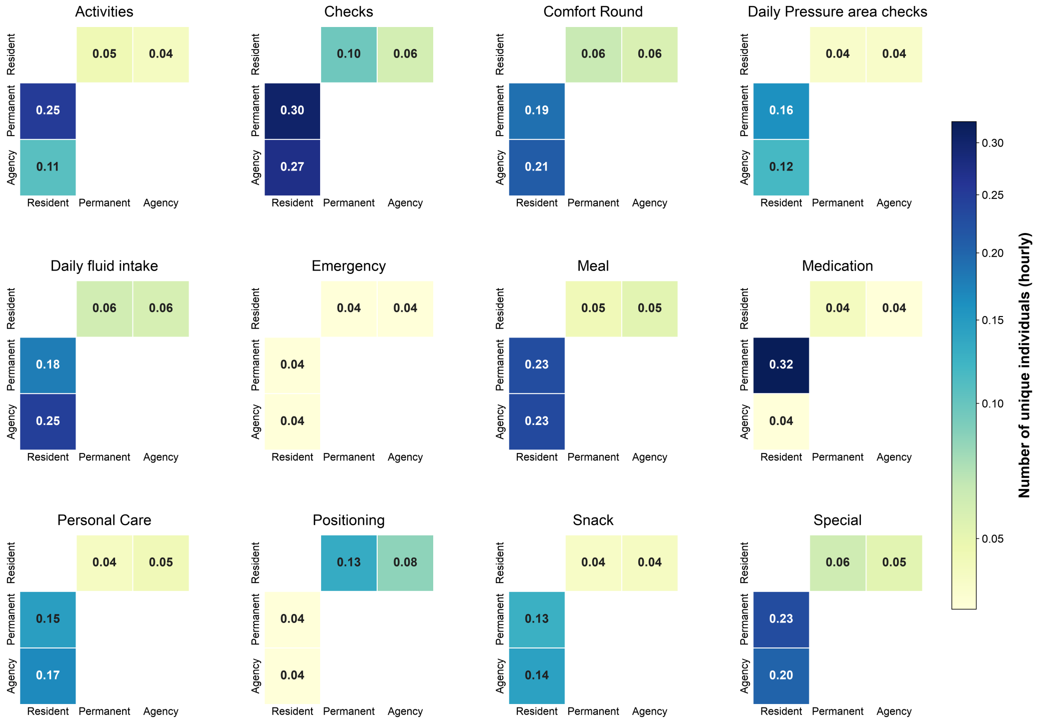


Figure S4: Service-specific hourly contact matrices using the number of unique individuals contacted as the contact definition. For each service category, matrices were constructed between residents, permanent staff, and agency staff using the processed care service records. Matrix cells represent the mean number of unique individuals contacted per hour, averaged over the observation period and normalised by time at risk. Colour intensity indicates the hourly mean, with darker blue representing more unique individuals contacted.

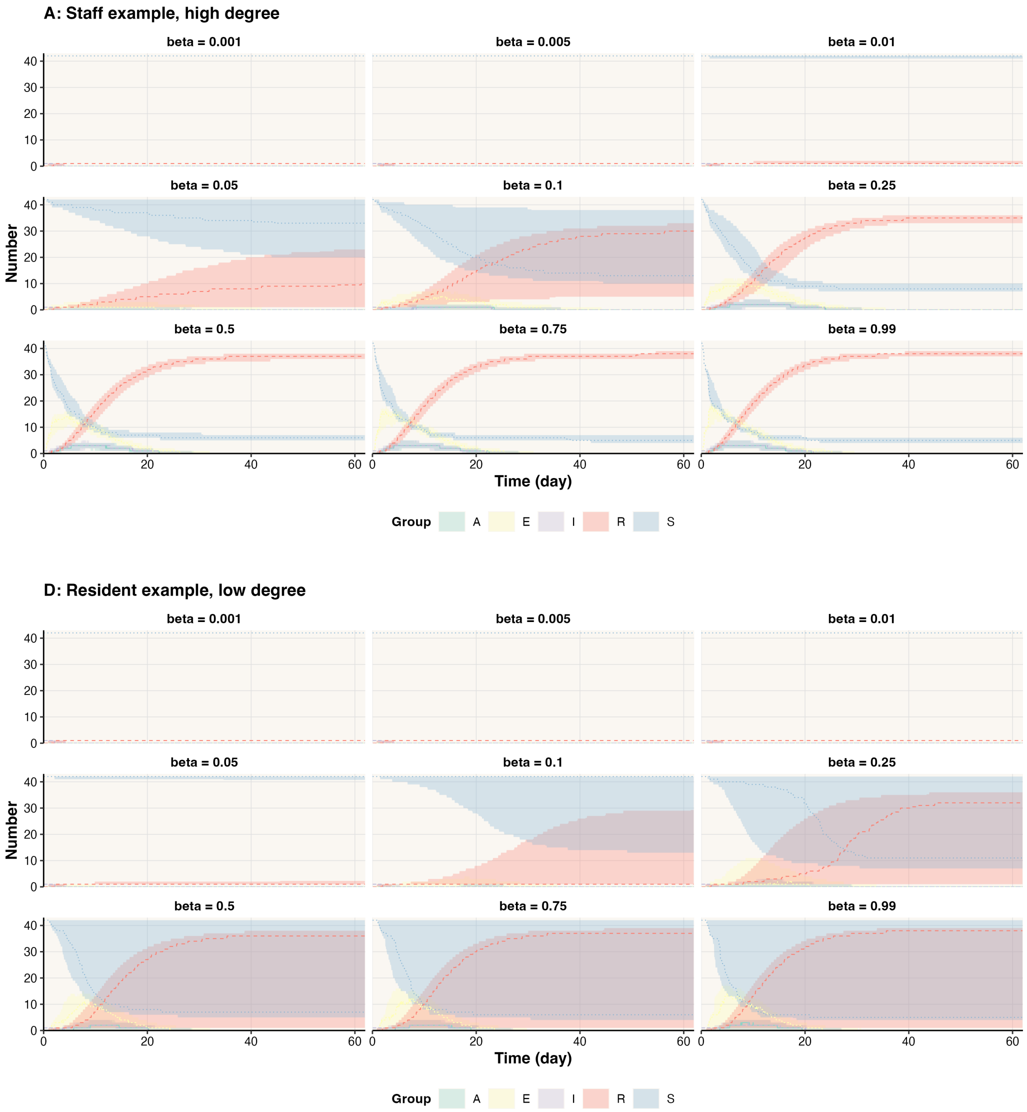


Figure S5: Temporal epidemic dynamics for the five epidemiological compartments (A, E, I, R, and S) across all nine transmission probabilities per contact ($\beta$), shown for simulations seeded by A, a staff example with high degree (top), and D, a resident example with low degree (bottom). For each seed individual and transmission probability, 1,000 simulations were performed. Lines show the median number of individuals in each compartment at each time point, and shaded bands show the interquartile range (25th–75th percentiles).

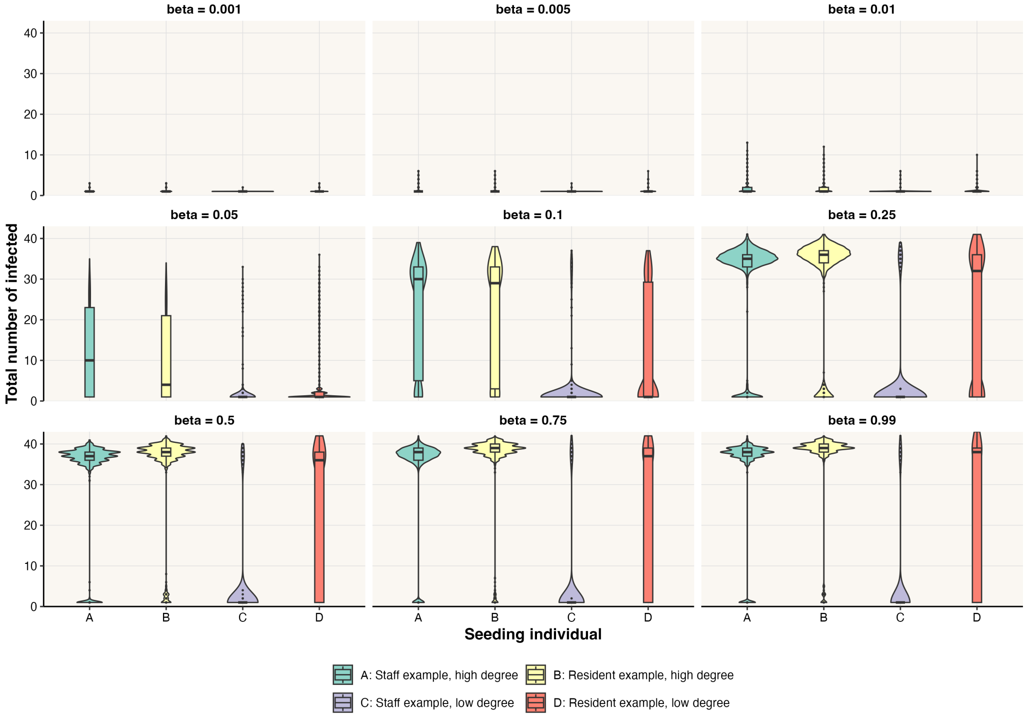


Figure S6: Distribution of the total number of individuals infected by the end of each simulation (final size) across all nine transmission probabilities per contact ($\beta$). Violin plots and embedded boxplots are shown separately for four seed individuals: A, staff example with high degree; B, resident example with high degree; C, staff example with low degree; and D, resident example with low degree. For each seed individual and transmission probability, 1,000 simulations were performed.

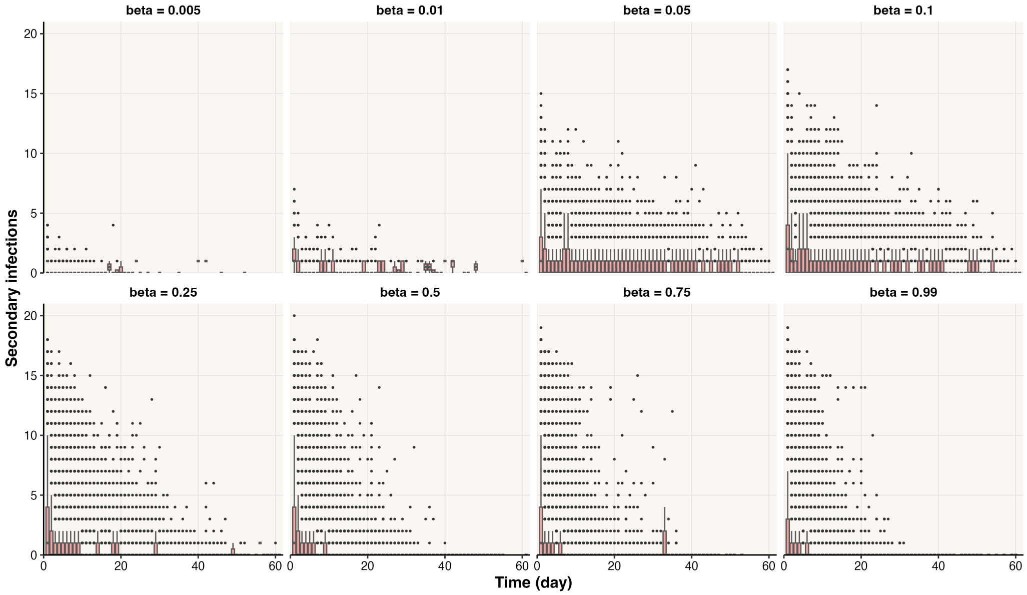


Figure S7: Distribution of the number of secondary infections caused by each infected individual, according to the day on which that individual became infected. For each transmission probability, data were pooled across the four selected seed individuals, with 1,000 simulations per seed. Results are shown for the eight transmission probabilities per contact ($\beta$ = 0.005, 0.01, 0.05, 0.1, 0.25, 0.5, 0.75, and 0.99) covered by the saved secondary-transmission records.

| Activity | Pattern Summary |
|---|---|
| Shift change | Day shift starts at 8am, and night shift starts at 8pm daily. |
| | Conduct start/end of shift check at the beginning and end of each shift. |
| Comfort round | Predominantly occurs when shift starts (morning and evening). |
| | Averagely each resident received 2.63 comfort rounds per day. |
| | More likely to happen in the morning but may occur sporadically during the day. |
| Medication | Administered at midday (11 am) and bedtime(7/8pm). |
| Fluid intake | Happens throughout the day and night, often coinciding with meal times |
| | Averagely each resident received 4.33 fluid intakes per day. |
| Activities | Limited to the day shift. |
| | Averagely each resident conducted 1.92 activities per day. |
| | Provided more by permanent staff compared to agency staff. |
| Food | Three meals were served everday for each resident, with breakfast at around 10am, lunch at around 2pm, and supper at around 7/8pm. |
| | Snacks were served during afternoons if needed. |
| Night check | Conducted approximately every 2 hours during night shifts. |
| Special check | Includes daily pressure area checks, emergency protocols, and other specific checks based on the residents' medical conditions. |

Table S2: Observed repeated service provision patterns

| **Metric** | **Daily weighted networks (n=61)** | | **Network for the whole duration (n=1)** |
|---|---|---|---|
| | **Mean** | **Standard Deviation** | |
| No. of active nodes | 25.93 | 2.55 | 43 |
| No. of links | 85.87 | 15.27 | 419 |
| Sum of weights | 211.31 | 38.45 | 13,101 |
| Density | 0.27 | 0.03 | 0.46 |
| Diameter | 3.88 | 0.45 | 3 |
| Average path length | 2.91 | 0.17 | 8.20 |
| Maximum path length | 6.74 | 0.27 | 33 |
| **Degree centrality** | | | |
| Mean | 0.27 | 0.03 | 0.46 |
| Maximum | 0.65 | 0.07 | 0.60 |
| Minimum | 0.07 | 0.04 | 0.12 |
| **Betweenness centrality** | | | |
| Mean | 0.05 | 0.01 | 0.05 |
| Maximum | 0.37 | 0.12 | 0.40 |
| Minimum | 0 | 0 | 0 |
| **Eigenvector centrality** | | | |
| Mean | 0.17 | 0.02 | 0.12 |
| Maximum | 0.47 | 0.07 | 0.32 |
| Minimum | 0.02 | 0.02 | 0.01 |
| **Strength** | | | |
| Mean | 16.33 | 2.79 | 609 |
| Maximum | 50.30 | 14.17 | 1750 |
| Minimum | 2.40 | 2.21 | 32 |

Table S3: Network metrics for daily weighted networks (n=61) and the aggregate network over the entire study period. Metrics include basic network properties (nodes, links, density) and centrality measures (degree, betweenness, eigenvector, and strength). For daily networks, mean and standard deviation are reported.
